# Untargeted longitudinal ultra deep metagenomic sequencing of wastewater provides a comprehensive readout of expected and unexpected viral pathogens

**DOI:** 10.1101/2025.10.27.25338874

**Authors:** Clayton Rushford, Devon Gregory, Emma Copen, Jonathan Naydenov, Alejandro Tovar-Mendez, Po-E Li, Rose S. Kantor, Migun Shakya, Nelson Ruth, Patrick S. G. Chain, Hsinyeh Hsieh, Torin Hunter, Josh Kome, Matt Frank, Terri Lyddon, Jeff Kaufman, David H. O’Connor, Marc C. Johnson

**Affiliations:** University of Missouri, Department of Veterinary Pathobiology; University of Missouri, Department of Molecular Microbiology and Immunology; B-GEN, Bioscience Division, Los Alamos National Laboratory; Physical and Life Sciences Directorate, Lawrence Livermore National Laboratory; University of Missouri, School of Natural Resources; Nucleic Acid Observatory, SecureBio; University of Wisconsin-Madison, Department of Pathology and Laboratory Medicine

**Keywords:** Wastewater surveillance, metagenomic, deep sequencing, SARS-CoV-2, influenza H5N1, influenza C, tomato brown rugose fruit virus, *NVD*, *GOTTCHA2*, One Health, public health

## Abstract

Wastewater surveillance has become a powerful tool to monitor circulating viruses at a community level. Currently, most wastewater surveillance efforts use target-based approaches such as quantitative PCR techniques or hybrid capture. This study explores the feasibility of using untargeted ultra-deep metagenomic sequencing as a comprehensive approach to wastewater surveillance. To test this, composite influent wastewater samples were collected weekly at a single site from January 2024 through June 2025. Sequencing was performed using random hexamers on all samples, with an average depth of 1.1 billion reads per sample. Human enteric viruses such as rotaviruses, astroviruses, and noroviruses were detected at high levels in virtually every sample. SARS-CoV-2 was also detected in most samples and the counts per sample positively correlated with digital PCR (dPCR) measurements. Less abundant respiratory pathogens such as influenza A and B, rhinoviruses, parainfluenzaviruses, and human coronaviruses 229E, OC43, NL63, and HKU1 were also regularly detected. However, those pathogens displayed distinct and reproducible winter and spring seasonality. Several unexpected viruses were also detected, such as several detections of highly pathogenic avian influenza H5N1 (HPAI H5N1) genotype B3.13, a month-long surge of hepatitis A virus, and a large season-specific surge in influenza C virus. The most abundant known virus detected was the *Tobamovirus* tomato brown rugose fruit virus, which was present stably year-round at high abundance. However, other tobamoviruses such as tomato mosaic virus were detected primarily in the late growing season. This eighteen-month study highlights that ultra deep sequencing enables detection of expected and unexpected viral pathogens without targeted enrichment.

**Importance:** This study demonstrates that untargeted ultra deep metagenomic sequencing can provide a comprehensive tool for wastewater surveillance of viral pathogens. By generating approximately 1 billion reads per sample across 78 consecutive weeks, we captured expected pathogens such as SARS-CoV-2, noroviruses, and influenza viruses. Additionally, we captured unexpected viral signals such as influenza C and highly pathogenic avian influenza H5N1. The wide range of viral taxa captured in this study also displays epidemiologically relevant seasonality. We also observed a correlation between metagenomic SARS-CoV-2 read counts and dPCR values to validate this method against other wastewater surveillance methods currently in use. Our findings highlight how ultra deep metagenomics can enhance pandemic preparedness, enable early detection of non-standard and clinically overlooked species, and broaden the scope of One Health monitoring by capturing human, animal, and plant viral signatures from a composite wastewater sample.

## Introduction

Wastewater surveillance is a powerful tool for public health that leverages minimally biased, population-level sampling, and supports pandemic preparedness by providing early warnings for circulating pathogens based on sequence data [1, 2]. Compared to clinical surveillance, wastewater-based approaches reduce biases associated with asymptomatic infections and limited access to healthcare. Several labs have demonstrated this concept through reverse transcription PCR (RT-PCR), real time quantitative PCR (RT-qPCR), and digital PCR (dPCR) designed to target human-infecting pathogens such as SARS-CoV-2, influenza, and *Enterovirus* spp. [3–6]. However, PCR-based approaches require occasional modifications due to evolution around primer binding sites and are limited to known targets [7]. Alternatively, probe-capture approaches facilitate detection of a wider range of pathogens in wastewater, have potential to detect pathogens that differ by up to 40% from reference genomes, and are commercially available [8–14]. While PCR and probe-capture approaches facilitate detection of viruses with *a priori* knowledge of genetic makeup, they are unable to detect highly divergent, novel viral sequences that may be circulating in a population.

Untargeted sequencing and metagenomics enable detection of a broader diversity of known and unknown pathogens if they receive sufficient coverage. Pathogen concentration methods, such as polyethylene glycol (PEG) precipitation and vacuum-based direct capture, have been shown to be the most reproducible while improving depth and breadth of viral signals [15]. However, a deep dataset is a prerequisite for untargeted metagenomic surveillance due to a low target-to-nontarget ratio of sequences [16]. Fortunately, the rapidly decreasing cost of sequencing has facilitated data generation that would, until recently, have been infeasible [17].

The goal of this study was to test the feasibility of viral surveillance using minimally biased metagenomic sequencing. A variety of previous studies have utilized metagenomic approaches for screening antimicrobial resistance genes, microbiomes, and viromes in wastewater [2, 18–26]. Most of these take place over a short period, yet a few longitudinal studies have surveyed wastewater for longer than a year to determine how detectable sequences change over time [11, 18, 27]. While previous studies have demonstrated the efficacy of metagenomics for viral surveillance, our study is unique due to our combination of longitudinal sampling (78 consecutive weeks), use of PEG precipitation for viral concentration, and sequencing at a depth greater than previous work [2, 11, 18–27]. To provide a high-confidence multi-kingdom readout of present species, we assigned sequence reads through *GOTTCHA2* (Genomic Origin Through Taxonomic CHAllenge) classification using an expanded reference database. For the detection of human-infecting viral sequences, we implement *NVD* (New Virus Discovery), a workflow designed to assemble putative viral sequences from human-infecting virus families and validate assemblies to be used as references for mapping abundance values.

## Methods

### Sample collection

Influent wastewater from Columbia, MO was collected weekly as a 24-hour composite sample using an ISCO autosampler. A 35 mL aliquot was transferred to sterile 50 mL conical and stored at 4°C prior to processing. At the time of collection, water quality parameters including temperature, pH, total suspended solids, average influent flow over sampling period, and chemical oxygen demand were recorded and provided by the Missouri’s Department of Health and Senior Services. A 35 mL aliquot of nuclease free water was used as a negative control for each batch of samples processed after October 8^th^, 2024.

### Viral concentration and RNA extraction for Metagenomics

Samples were centrifuged for 2000 xg at 4°C for 5 minutes and the supernatant was filtered through 0.22 µm filters (Millipore, SCGP00525) to remove solids and larger microbes. A subset of samples collected between January 2^nd^ and March 19^th^, 2024 were then treated with 10,000 ng of RNAse A (ThermoFisher, R1253), 2 units of TURBO DNAse (Invitrogen, AM1907), and a 4.75 mL of 5 mM CaCl_2_ and 25 mM MgCl_2_ buffer, in an attempt to remove bacterial ribosomal RNA. Separately, another sample subset was treated with nucleases (as above) and 30 mM EDTA to dissociate rRNA from ribosomes. For nuclease treated samples, 400 units of RNaseOUT™ Recombinant Ribonuclease Inhibitor (Invitrogen, 10777019) was added prior to RNA extraction. A breakdown of treatments per sample date is summarized in Supplemental Table 1.

Untreated, nuclease treated, and nuclease and EDTA treated sample filtrates were mixed with 12.5 mL of 50% (w/vol) PEG (Research Products International, P48080-1000.0) and 1.2M NaCl for 1 h at 4°C. Nucleic acids were extracted from pellets using QIAamp Viral RNA Mini Kit (Qiagen, 52906) using the manufacturer’s instructions excluding the addition of carrier RNA. Samples were not DNase treated after extraction.

### RT-qPCR of pepper mild mottle virus

Viral RNA in samples was assessed prior to library preparation through comparative RT-qPCR targeting pepper mild mottle virus (PMMoV) to determine if viruses were successfully concentrated. Pepper mild mottle virus was selected as an endogenous control due to its ubiquitous distribution in wastewater [28]. Forward (5’ GGCGTAGATCCATTGGTGC 3’) and reverse (5’ CGAACCTTCCTCCTTTGATG 3’) primers were designed for RT-qPCR to target the PMMoV genome were mixed and resuspended at 100 µM concentrations. A Taqman probe (VIC-5’ GCTGTGGTTTCAAATGAGAGTGG 3’-QSY) was prepared at 100 μM concentration. The RT-qPCR reaction was carried out using TaqPath 1-Step RT-qPCR Master Mix, CG (4x) (Applied Biosystems, A15299), forward and reverse primers (500 nM each), taqman probe (125 nM), and 5 μL of RNA template for a total reaction volume of 20 µL. Reactions were carried out on an Applied Biosystems 7500 Fast System, with thermocycler settings of 48°C for 15 min, 95°C for 10 min, followed by 40 cycles of 95°C for 15 s and 60°C for 1 min.

### Library preparation and deep sequencing

Total RNA concentrations were quantified through Qubit RNA High Sensitivity Assay, although some samples were below the detection limit for this assay. Libraries were prepped from RNA for deep sequencing using Illumina’s Stranded mRNA Prep, Ligation (Illumina, 20040534) and Illumina RNA UD Indexes Sets A-D (Illumina, 20091655, 20091657, 20091659, 20091661) following manufacturers recommendations with slight modifications listed below. RNA inputs for library preparation were normalized based on Qubit results to 10-15 ng in 9.5 µL. If input concentrations were <0.2 ng/μl then 9.5 µL was used for input. The mRNA capture step was omitted and library preps began with RNA fragmentation and cDNA synthesis. Normalized RNA inputs were evenly split into two aliquots. One aliquot was fragmented so longer RNAs were broken into a target range compatible with sequencing, while the other half was left unfragmented to retain sequences already within the target range. After fragmentation, aliquots were pooled together for cDNA synthesis. Lastly, final indexed libraries underwent size selection during the final cleanup by mixing 30 μL of AMPure XP (Beckman Coulter, A63881) with 50 μL of post-indexing PCR amplicons to bind to long library sequences. Magnetic beads were incubated on a magnetic stand for 5 minutes. The supernatant was separated from the bead pellet and moved to a new tube where 20 μL of AMPure XP beads were added to bind to shorter library sequences. These shorter fragments were washed, eluted in 30 μL of resuspension buffer, and used for sample pooling. Final libraries were sequenced using a NovaSeq 6000 until April 16^th^ 2024, and subsequent libraries were sequenced using a NovaSeq X with PE150 reads (Supplementary Table 1).

### Assessing sequencing depth and rRNA abundance in samples

Analysis of rRNA abundance (described below) and qPCR of PMMoV showed no noticeable deviation in percent rRNA or Ct values for our endogenous control in sample dates that underwent multiple treatments. Therefore, treatment use was discontinued, and fastq files from samples with multiple treatments per date were combined prior to taxonomic assignment. Fastq files were trimmed for adapters, homopolymers, low entropy sequences, and short length sequences using fastp (version 0.23.4) using the following options, “-w 16 --dont_eval_duplication -l 50 -r -c --trim_poly_g --poly_g_min_len=5 --trim_poly_x --poly_x_min_len=5 -y --adapter_sequence=CTGTCTCTTATACACATCT --adapter_sequence_r2=CTGTCTCTTATACACATCT”. Samples were split into rRNA and non-rRNA fractions to assess the approximate depth of potential viral sequences. Ribosomal RNA separation was performed using bbduk.sh from BBmap (version 38.18) and a custom rRNA database curated by Brian Bushnell [29]. The depth of rRNA fractions and non-rRNA fractions were determined using stats.sh from BBmap. Both BBTools commands were performed using default settings.

### Taxonomic assignment with GOTTCHA2

Overall sample composition was determined with high confidence through a species-level *GOTTCHA2* (version 2.1.9.0) analysis using fastp trimmed sequences and arguments “-nc -rm no” [30]. *GOTTCHA2* was selected for reporting for definitive species presence due to its curated multi-kingdom reference database which uses unique genomic signatures at the queried taxonomic level. Using the arguments listed above, the detection threshold for this analysis requires sequences to have a minimum linear coverage of 0.005, minimum of 3 reads, minimum length of 60 bases, maximum estimated z-score of 30 for the depth of mapped region, and a minimum mapped fraction of 50% to the signature fragment.

Taxonomic assignments were grouped by species, read counts were summed for each species, and read counts were normalized to reads per million (RPM). Host domains for each detected viral phylum were manually annotated by searching host range of the detected phylum. Sources and reasoning for annotations are listed in Supplemental Table 2. To ensure that community composition derived from *GOTTCHA2* is consistent with a more permissive classifier, *Kraken2* was also used for taxonomic assignment. *Kraken2* (version 2.1.3) was run on trimmed R1 and R2 files using the arguments “--use-names --paired” and the PlusPFP database (accessed June 2024). *Kraken2* report read counts were normalized to RPM and only reads classified on the species level were used for comparison with *GOTTCHA2.* Unclassified reads from *GOTTCHA2* and *Kraken2* were not included in outputs.

### Taxonomic assignment with NVD

We developed a Snakemake-based pipeline to detect and classify viral sequences from high-throughput sequencing data. The pipeline was containerized using Apptainer (formerly Singularity) to ensure reproducibility across computing environments. Raw sequencing reads were first interleaved using BBMap (v38.18). To reduce computational requirements and improve specificity, reads were pre-filtered using NCBI’s Sequence Taxonomy Analysis Tool (STAT; aligns_to v3.1.1) to retain only those with k-mer matches to known human-infecting virus families, including *Adenoviridae*, *Anelloviridae*, *Arenaviridae*, *Astroviridae*, *Bornaviridae*, *Peribunyaviridae*, *Caliciviridae*, *Coronaviridae*, *Filoviridae*, *Flaviviridae*, *Hepadnaviridae*, *Hepeviridae*, *Orthoherpesviridae*, *Orthomyxoviridae*, *Papillomaviridae*, *Paramyxoviridae*, *Parvoviridae*, *Picobirnaviridae*, *Picornaviridae*, *Pneumoviridae*, *Polyomaviridae*, *Poxviridae*, *Sedoreoviridae*, *Retroviridae*, *Rhabdoviridae*, *Togaviridae*, and *Kolmioviridae*. *Arteriviridae* were also included due to their ability to infect other primate species. Unclassified reads at this step were discarded.

The filtered reads were assembled using SPAdes (v4.0.0) in sewage mode. Full genome reconstruction was not a goal of this study, but generation of assemblies would create a reference database to be used for subsequent abundance mapping. The resulting contigs underwent quality control using BBMask to remove low-complexity regions using an entropy threshold of 0.9. Following masking, contigs were filtered to retain only sequences with at least 200 consecutive non-N bases after trimming terminal N’s, as shorter sequences can interfere with subsequent classification steps. The quality-controlled contigs then underwent a hierarchical classification process beginning with analysis using a NCBI STAT’s tree index database provided by Kenneth Katz (downloaded from NCBI on 2024-08-30; tree_filter.dbss, tree_filter.dbss.annotation, tree_index.dbs). From this first-pass classification, a focused taxa list was generated and used for a second, more detailed classification using STAT’s dbss database. This two-stage STAT analysis used a STAT cutoff-percent value of 0.001% (the default is 0,01) to more sensitively include matching contigs for subsequent BLAST analysis.

The BLAST-based classification employed a two-stage approach for maximum sensitivity. Initially, contigs were analyzed using MEGABLAST against NCBI’s nucleotide database with default parameters to identify close matches to known sequences. Contigs that remained unclassified after MEGABLAST underwent a more sensitive BLASTN search using default parameters. The results from both BLAST stages were filtered to remove non-viral matches and bacteriophages, then merged to create a comprehensive classification of the viral contigs. The workflow does not enforce explicit detection thresholds for alignment length, percent identity, coverage, or E-value, but instead relies on BLAST defaults and returning the top 5 hits per contig and selects a single hit based on the lowest E-value. To validate the assembled viral sequences and assess their coverage, the initially filtered reads were mapped back to the identified viral contigs with minimap2 using the options “-ax sr” (v2.1). Coverage calculations were performed using samtools (v1.20), retaining only primary alignments for accurate depth estimation. A SQL script was generated to query BLAST validated assemblies for human specific pathogen names and group them based on tropism for use in heatmap generation. Similarly to *GOTTCHA2* read counts, *NVD* read counts were normalized to RPM for each species detected.

*NVD* derived influenza H5N1 assemblies were aligned for visualization using minimap2 on default settings with Influenza A virus (A/Grackle/Texas/USL_047/2024(H5N1)) as a reference genome because 5 of the 9 assembled contigs were blast validated as this strain. Next, samtools was used to filter out quality scores less than 30 and secondary alignments and index the alignment for viewing with IGV (version 2.16.2). Influenza C assemblies were also aligned using the same method and genome coverage over the course of the study was assessed with samtools using the option “depth -aa”. The most abundantly detected influenza C strain, influenza C virus (C/Sri Lanka/68/2023), was used for reference.

### Digital PCR targeting SARS-CoV-2

Raw samples were prepped in accordance with Missouri’s SARS-CoV-2 wastewater surveillance program. Twenty-four-hour composite samples as described above were acquired at Missouri wastewater treatment facilities (WWTF) and stored at 4°C until processing. After mixing the samples briefly by gently inverting the tubes, a total of 9.75 mL of wastewater per sample was loaded into 24-deep-well plates. A total of 20 μL of the previously described NL4-3 derived HIV with the CMV-driven puromycin-resistance gene and lacking the *env*, *nef*, *vif*, and *vpr* genes were added to each sample as a processing control [31]. Then, a total of 100 μL of Nanotrap Enhancement Reagent 2 (Ceres Nanosciences, 10112-10) and 150 μL of Nanotrap Microbiome A Particles (Ceres Nanosciences, 44202) were added to each sample to capture, concentrate, and enhance downstream detection of viruses. A total of 5.6 μL of carrier RNA (Qiagen, 52906) was mixed with 700 μL of Buffer AVL (Qiagen, 52906) on a separate 24-deep-well plate to lyse viruses and help with RNA recovery from the wastewater samples. All plates were loaded into the KingFisher Apex (ThermoFisher Scientific) and processed. After processing in the KingFisher Apex, 700 μL of the lysate was loaded into the QIAcube Connect (Qiagen), and RNA was extracted and eluted into a final volume of 60 μL using the QIAamp Viral RNA Mini Kit (Qiagen, 52906), according to the manufacturer’s instructions.

Forward (5’ GACCCCAAAATCAGCGAAAT 3’ and 5’ TTACAAACATTGGCCGCAAA 3’) and reverse (5’ TCTGGTTACTGCCAGTTGAATCTG 3’ and 5’ GCGCGACATTCCGAAGAA 3’) primers and TaqMan probes (FAM-5’ ACCCCGCATTACGTTTGGTGGACC 3’-MGB-NFQ and ABY-5’ ACAATTTGCCCCCAGCGCTTCAG 3’-QSY) were designed for RT-dPCR to target the N1 and N2 regions of N gene of SARS-CoV-2, respectively. The primers were resuspended at 100 μM concentrations as TaqMan probe. The RT-dPCR reaction was carried out using QIAcuity One Step Advanced Probe kits (Qiagen, 250131), forward and reverse primers (500nM each), TaqMan probe (125 nM) and 10 μL of RNA template for a total reaction volume of 40 μL in a QIAcuity Four digital PCR system, with thermocycler settings of 50°C for 30 min, 95°C for 2 min, followed by 45 cycles of 95°C for 10 s and 55°C for 30 s, after priming.

### Statistical analysis

Relative abundance of rRNA was assessed for correlation against collection temperature, water pH, total suspended solids, average influent flow over sampling period, and chemical oxygen demand using Spearman’s Correlation Coefficient.

For *GOTTCHA2* assigned sequences, Shannon–Wiener diversity (H′) at the species level was calculated separately for prokaryotic-host and eukaryotic-host viruses in each sample using the diversity function in the Vegan package (version 2.6.8) of R (version 4.4.2). Paired differences in H′ between prokaryote-infecting and eukaryote-infecting viruses were tested for normality by Shapiro–Wilk test, then compared using a two-tailed paired Student’s t-test. A Wilcoxon signed-rank test was also performed as a nonparametric alternative. A 95% confidence interval for the mean H′ difference was estimated by nonparametric bootstrap (10,000 replicates) via the boot package (v1.3-31). Statistical significance was defined as p < 0.05.

Metagenomic read counts for SARS-CoV-2 were compared with dPCR reports through Missouri’s SARS-CoV-2 wastewater surveillance program to determine if the metagenomic approach showed consistent epidemiological trends and was consistent with our current SARS-CoV-2 surveillance protocol. For each sampling week, dPCR results were flow normalized by multiplying mean copies per liter by average influent flow during the sampling period to calculate total SARS-CoV-2 viral load. The metagenomic SARS-CoV-2 (NC_045512.2), PMMoV (NC_003630.1) and ToBRFV (KT383474.1) sequences were extracted from trimmed fastq files using minimap2 (version 2.14-r883) using the argument “-ax sr”. Secondary alignments and matches with MAPQ scores <30 were removed using samtools (version 1.20). The association between flow-normalized dPCR or with SARS-CoV-2 counts normalized to RPM, PMMoV, or ToBRFV were quantified in R using Pearson’s correlation (two-sided). For visualization only, the metagenomic counts were linearly rescaled to overlay the dPCR results on a secondary y-axis.

For *NVD* assigned sequences and *GOTTCHA2* sequences assigned to the same viral families, beta diversity was longitudinally assessed on the species level through Bray-Curtis dissimilarity of both outputs using the vegdist function from the vegan package in R. Permutational analysis of variance (PERMANOVA) was performed to evaluate the effects of season on the full data set. Since samples were collected longitudinally from a single site, results were interpreted as indicative of overall compositional differences rather than as strictly independent statistical tests, recognizing the potential for temporal autocorrelation among samples. Seasons were assigned based on solstice and equinox dates. PERMANOVA was also used to evaluate season, year, and their interaction on a subset of data focusing on winter and spring. Non-Metric Multidimensional Scaling (NMDS) was performed with *k* = 2 dimensions and ordination was repeated with multiple random starts (up to 200 iterations) to ensure stable solutions. The first two NMDS axes were plotted to visualize community dissimilarity.

## Results

### Sequencing depth and sample quality

Raw 24-hour composite Influent wastewater samples from Columbia, MO were collected weekly from January 2024 through June 2025. Samples were filtered, PEG concentrated, and extracted nucleic acids were sequenced using untargeted ultra-deep sequencing. While this study targeted RNA viruses due to the method used for library preparation, purified nucleic acids contained both RNA and DNA, and DNA viruses were included in the output. Over the study period, 85.8 billion read pairs were obtained with an average depth of 1.1 billion read pairs per sample (**Figure 1**). On average, ribosomal RNA (rRNA) accounted for 66.2 ± 20.7% of reads per sample (range 14.2% - 95.5%). No strong correlations were observed (|ρ| ≥ 0.5) between rRNA abundance and temperature, water pH, total suspended solids, average influent flow rate over the sampling period, or chemical oxygen demand associated with samples. However, high rRNA abundances are expected in nonselective metagenomic sample preps [21, 22]. The average RT-qPCR Ct value for the wastewater-ubiquitous plant virus PMMoV was 24.2 ± 1.1, providing evidence of viral concentration, even when overall RNA concentrations were low or below the detection limit. Negative control samples were prepped on a per-batch basis starting on October 8^th^ 2024 and used to rule out contamination of suspicious hits when assessing taxonomic profiles.

**Figure 1.**
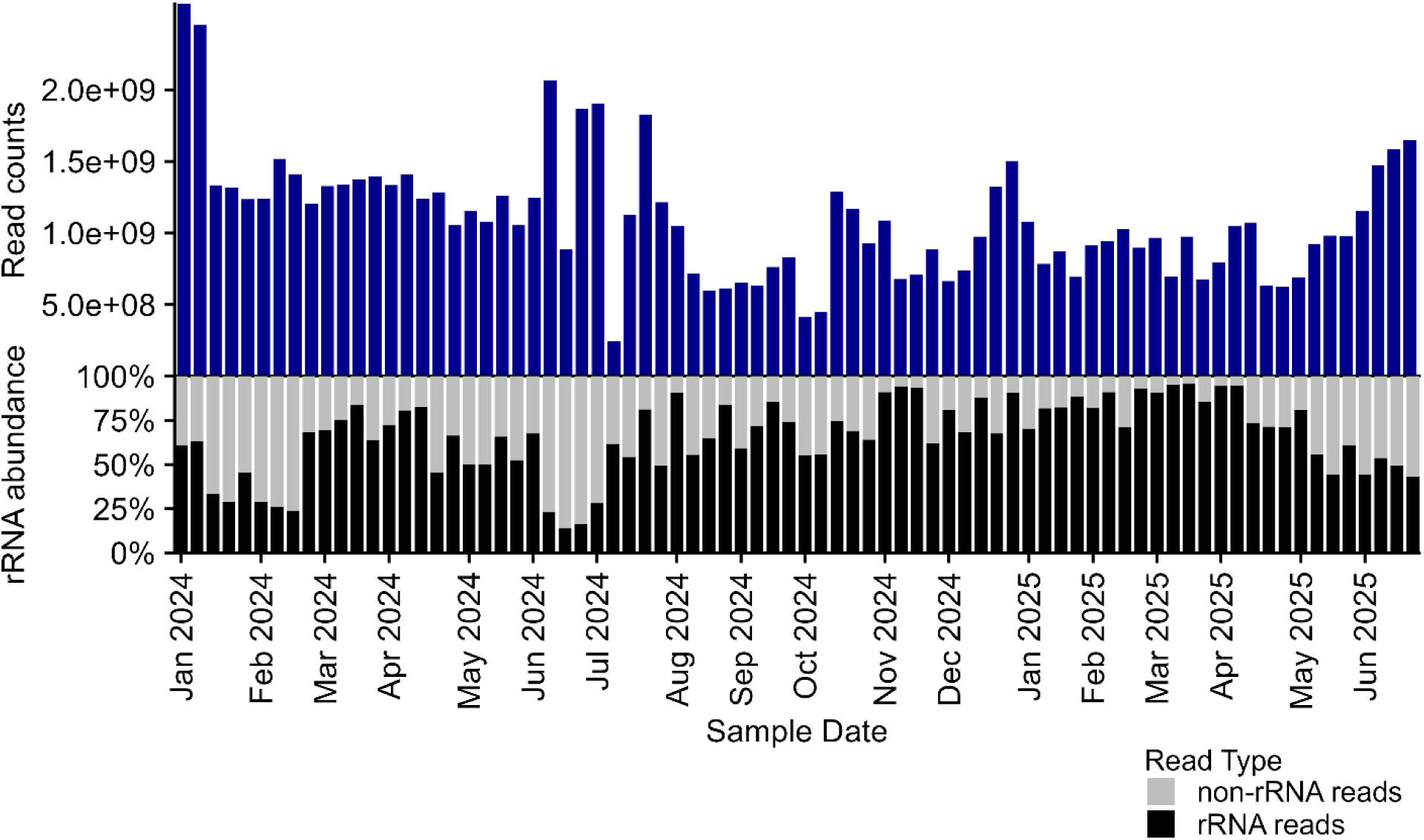
Sequencing depth and rRNA abundance of Columbia MO wastewater samples from January 2024 through June 2025. Blue bars represent total read counts for each sample date. The x-axis indicates the first sample date of the corresponding month labeled.

### Definitive taxonomic profile

We used *GOTTCHA2* to identify reads that matched unique genomic regions specific to each taxonomic level. The objective of this classification method was not to provide a readout of total sample composition, but provide a baseline for species that are unambiguously present in each sample. The settings used for this analysis enforce a stringent mapping threshold, ensuring that a read must match at least 30 bp of a signature fragment. The average percentage of sequences that could be assigned at the species level using *GOTTCHA2* was 3.79% ± 1.78% representing approximately 40 million reads per sample. In nearly all samples, eukaryotic viruses were more abundant than prokaryotic viruses (**Figure 2A**). The most abundant viral family, *Virgaviridae*, consists mostly of tomato brown rugose fruit virus (ToBRFV), with PMMoV also consistently present throughout the year but at lower levels (**Figures 2B and 2D**). Notably, other plant viruses such as tomato mosaic virus (TMV) and cucumber green mottle mosaic virus (CGMMV) were detected predominantly during US growing seasons (**Figure 2D**). A similar breakdown of abundant prokaryotic virus families is shown in **Supplemental Figure 1**. Taxonomic assignment using *Kraken2* generated community profiles nearly identical taxonomic distribution and the same highly abundant viral families as *GOTTCHA2* (Supplemental Figure 2 and 3).

**Figure 2.**
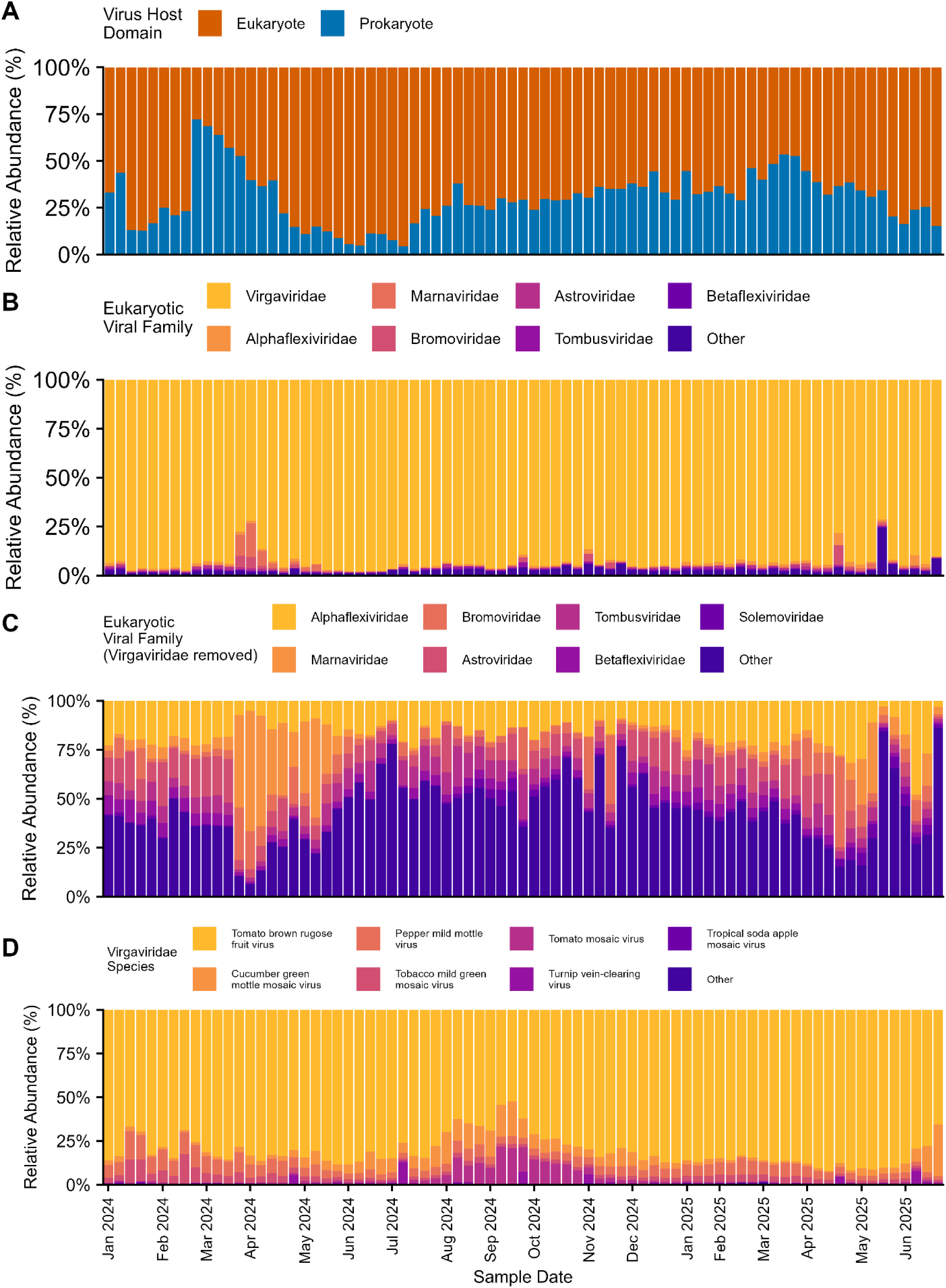
Normalized abundance (RPM) of *GOTTCHA2* classified viral reads out of total classified viral reads, colored by host domain based on manual annotation (A). Normalized abundance (RPM) of the seven most abundant eukaryotic infecting virus families (B). Normalized abundance (RPM) of the seven most abundant families within eukaryotic viruses after removal of the family *Virgaviridae* (C). Breakdown of the most abundant species within the family *Virgaviridae* (D).

**Figure 3.**
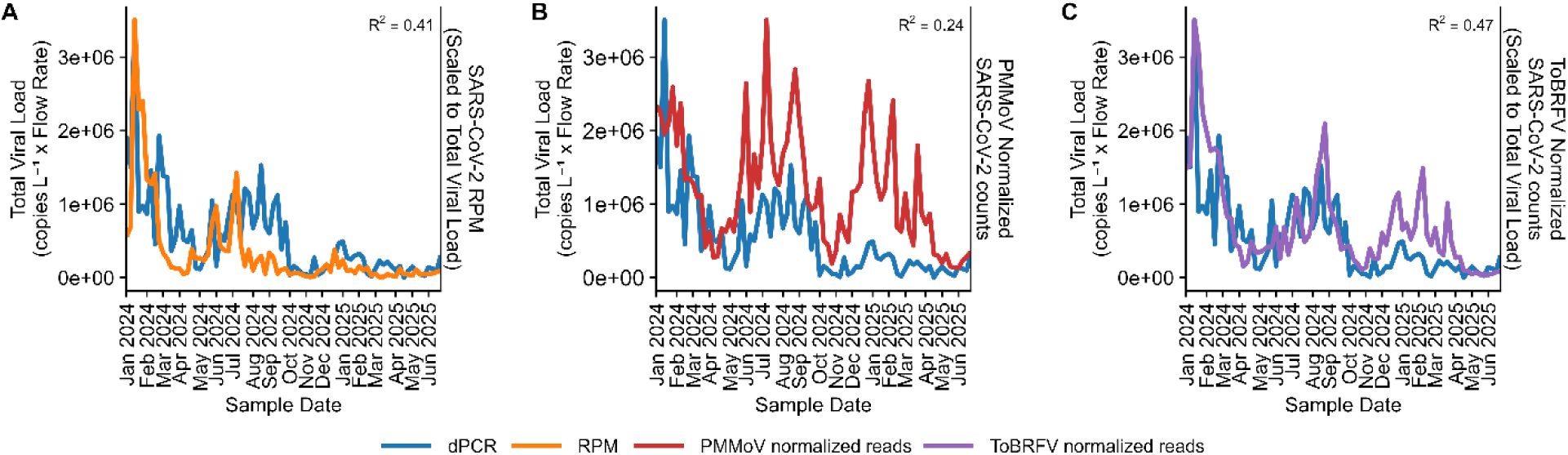
Comparison of SARS-CoV-2 read counts derived from metagenomics using minimap2 against reference genomes and dPCR results from Missouri’s Wastewater Surveillance Program from January 2024 through June 2025. SARS-CoV-2 read counts normalized to RPM (A), PMMoV (B), and ToBRFV (C) were scaled prior to plotting by dividing the max value of clinical cases or total viral load by the max value of SARS-CoV-2 read counts.

Although prokaryotic viruses were less abundant in most samples, the prokaryotic virus fraction was much more diverse compared to the eukaryotic virus fraction (**Supplemental Figure 4**). Mean Shannon diversity was 3.15 ± 0.42 for prokaryotic viruses and 1.10 ± 0.24 for eukaryotic viruses with a paired mean difference of 2.05 (95% CI 1.94-2.16) that was statistically significant (paired t-test *p* < 2.2 × 10^−16^, Wilcoxon *p* = 2.51 × 10^−14^). If the distribution of species were even, then these values would roughly equate to prokaryotic virus fractions having 8-fold more detectable species than the eukaryotic virus fraction. However, this difference is primarily due to the strong ToBRFV signal skewing the diversity of the eukaryotic virus fraction, causing subsequent underreporting of eukaryotic viral diversity.

### Comparison between dPCR and metagenomic read counts

Many metagenomic studies normalize read counts by an endogenous control, such as PMMoV, so counts are more reflective of true viral signal trends and not skewed by changes in wastewater flow rate or variability in sample preparation. Since ToBRFV was the most abundant virus while also displaying the smallest coefficient of variance throughout the study of the highly abundant viruses such as PMMoV, we compared SARS-CoV-2 metagenomic counts normalized to RPM, PMMoV, and ToBRFV to dPCR findings from Missouri’s wastewater surveillance program. SARS-CoV-2 counts normalized to ToBRFV showed the best correlation with flow normalized dPCR measurements (p = 4.2e^−12^, R^2^ = 0.471) compared to RPM (p = 1.8e^−10^, R^2^ = 0.41) and PMMoV (p = 4.5e^−6^, R^2^ = 24; **Figure 3**).

### Human virus profiles

To better investigate human infecting viruses, we developed and applied a custom pipeline to extract putative human infecting virus reads, assemble them into contigs for BLAST validation, and derive abundance values by mapping raw reads to assembled contigs. This pipeline facilitates detection of a wide breadth of pathogens that are non-standard in probe panels for wastewater or overlooked in clinical settings. Many viruses and viral groups were detected across the study period, such as SARS-CoV-2, enteroviruses, rhinoviruses, and parechovirus A (**Figure 4**) while other viruses and viral serotypes were only present at distinct seasons or timeframes (**Figure 5**). For example, during the winter and early spring months of both years, the classic respiratory seasonal viruses such as influenza A virus, and human coronaviruses NL63 and OC43 were readily detected, while during the summer months these viruses were virtually absent (**Figure 5**). Some respiratory viruses appeared to surge during specific seasons. For instance, in both 2024 and 2025 influenza A virus was predominantly detected from January to March, while parainfluenza virus 3 detection was primarily from March to June, which is consistent with what is observed clinically [32, 33]. Enterovirus D68, on the other hand, was detected primarily in the fall, consistent with clinical observations [34]. Some of the viruses detected had a more complex distribution when the specific serotypes were considered. For instance, rhinoviruses were detected year-round, with higher detection levels in the winter and spring (**Figure 4**). However, when individual serotypes were analyzed, there were distinct waves (**Figure 5**). While most rhinovirus serotypes were abundant in the late spring months (e.g. A58, A18, and A68 in the spring of 2024), some serotypes surged in the fall and early winter, such as rhinovirus C42 in late 2024. Notably, while rhinoviruses were most pronounced in the spring, the season-specific serotypes were not the same from year to year. For instance, rhinoviruses A61 and A30 were both prevalent in the spring of 2025, but were completely absent in the spring of 2024. Rhinovirus infections are rarely typed, so it is difficult to say if these observations are consistent with what was seen clinically.

**Figure 4.**
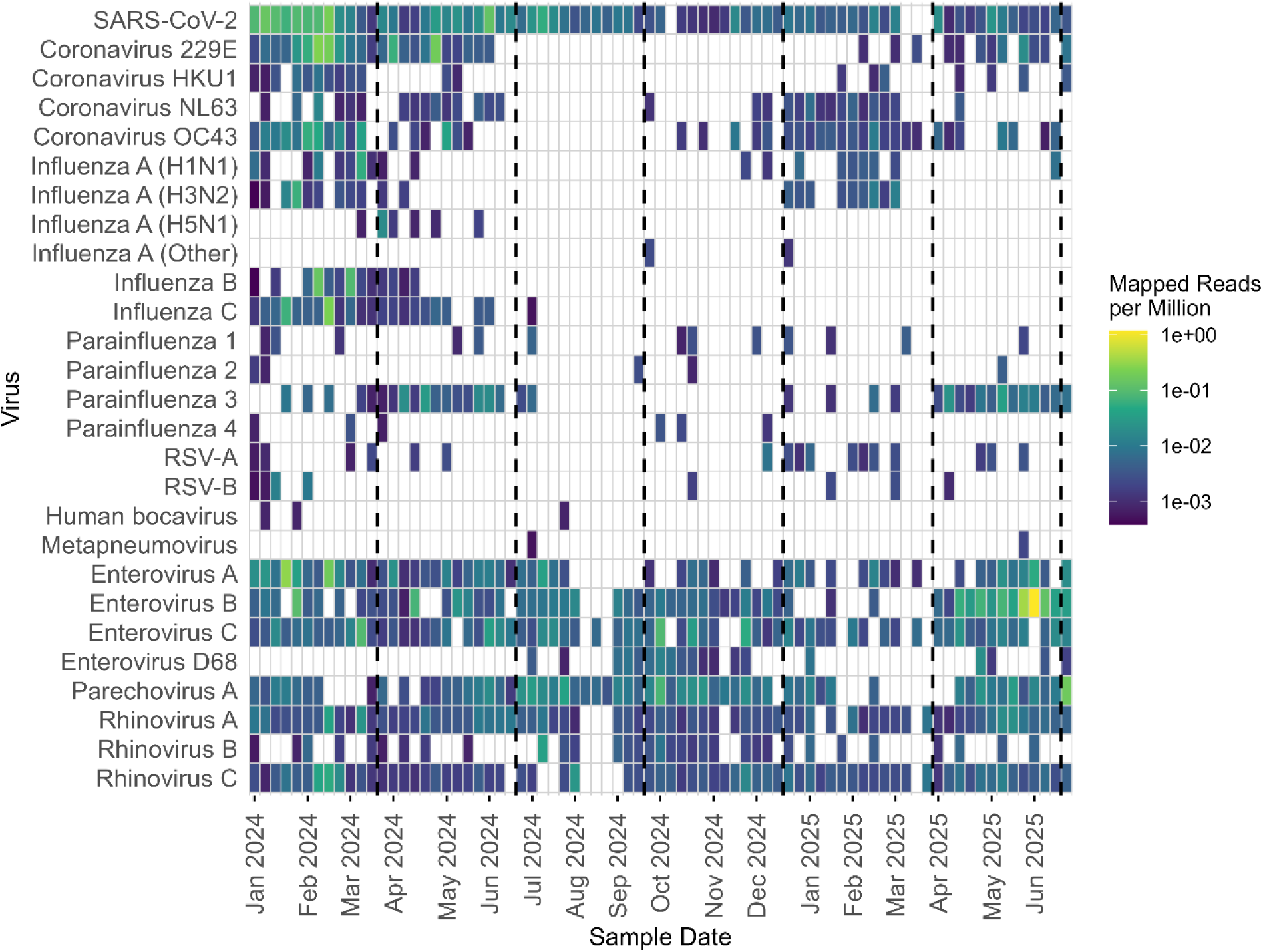
Normalized abundance (RPM) of human respiratory viruses in Columbia MO wastewater from January 2024 to June 2025. Reads counts were derived by mapping raw sequences to NVD assembled contigs and counts were normalized to mapped reads per million (RPM) prior to plotting as opposed to mapped reads per kilobase per million (RPKM) due to groupings of viruses with different genome lengths in some rows. A log_10_ color scale was used for visualization to enhance dynamic range and improve interpretability of variation across samples. Vertical dashed lines signify solstice and equinox dates to highlight season timeframes.

**Figure 5.**
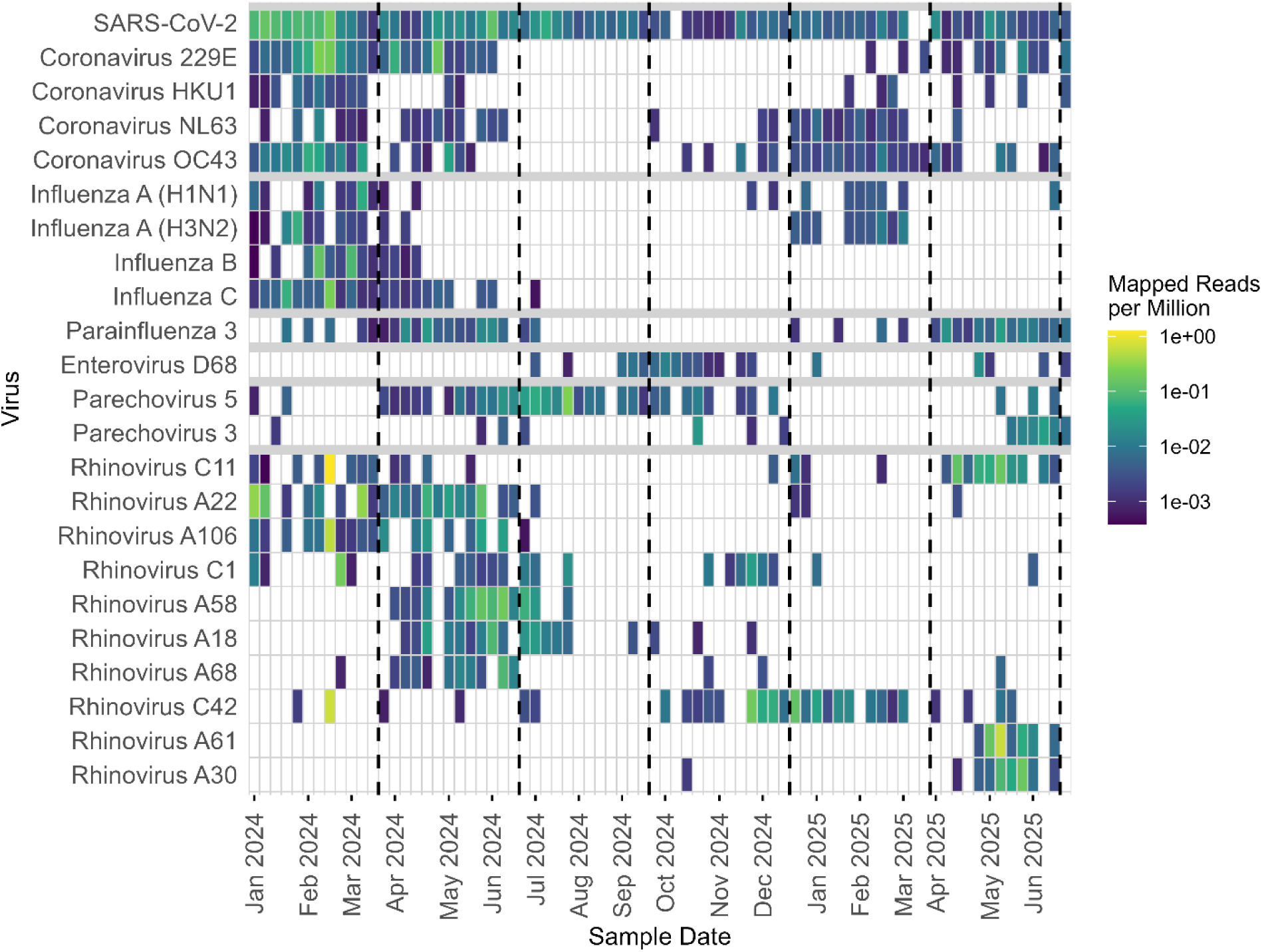
Normalized abundance (RPM) of individual *NVD* assigned viruses associated with human respiratory disease that exhibit a seasonal trend. Reads counts were derived by mapping raw sequences to NVD assembled contigs and counts were normalized to mapped reads per million prior to plotting to keep normalization consistent across heatmaps. A log_10_ color scale was used for visualization to enhance dynamic range and improve interpretability of variation across samples. Vertical dashed lines signify solstice and equinox dates to highlight season timeframes.

The parechoviruses also displayed a year-to-year serotype change. In 2024, parechovirus 5 was the main serotype detected in 2024, while parechovirus 3 was the primary serotype in 2025. This is consistent with a report that demonstrated that parechovirus 5 emerged as the dominant serotype in nearby Kansas City in 2024 [35].

Some of the human-infecting viruses detected were not expected. One example was detection of the respiratory pathogen influenza C virus, an influenza type that generally causes milder disease and is typically not included in respiratory panels (**Figures 4 and 5**). Influenza C virus was the most frequently detected species of influenza in the winter of 2023/24, but did not return the following winter. Coverage ranged from 31-100% across coding sequences, with all seven Influenza C genome segments represented (Supplemental Figure 5). Another unexpected finding from wastewater was the detection of influenza A virus subtype H5N1 from early March-May of 2024 (Supplemental Figure 6). The assembled sequence matched the influenza H5N1 strain B3.13 found in the mammary tissue of dairy cattle and reported by the USDA in late March 2024, followed by dissemination to 16 states by the end of the year [36]. Additionally, our H5N1 detections align to the same timeframe as detections throughout Texas wastewater [37]. While Missouri did not have any reported H5N1 dairy herds, the signal was believed to be derived from a local dairy that received milk for processing from a state with H5N1-infected herds. This finding highlights the fact that this metagenomic approach can detect pathogen surges that are likely to be missed by standard patient surveillance.

Viruses associated with gastrointestinal disease, such as norovirus genogroup II, norovirus genogroup I, rotavirus A, astroviruses, Sapporo virus, human mastadenovirus F, and human picobirnavirus were detected throughout the study (**Figure 6**). Some viruses, such as rotavirus A and astrovirus MLB, displayed slight increases during colder months. A month-long surge of hepatitis A virus was detected the summer of 2024, but the signal did not return for the remainder of this study. As expected for wastewater, read counts for enteric viruses were higher than detected respiratory viruses (**Figures 4-6**). Despite the lower counts associated with respiratory viruses, all reported human-infecting viruses were captured with sufficient coverage for raw reads to assemble with SPAdes, undergo BLAST-based classification, and read-mapping–based coverage assessment. When analyzed in terms of the relative abundance of viruses at the species level using only viral families listed in *NVD* analysis, season explained 18.8% of the variance of sample groupings for *NVD*, and 27.3% of the variance for *GOTTCHA2* (p < 0.001 for both methods; **Figure 7**). Additionally, both year and the interaction between season and year were significant (p < 0.01) for spring and winter suggesting that seasonal trends are mostly consistent between years with slight shifts in community composition from year to year. The seasonality associated with this data reflects known epidemiological patterns and reinforces previous studies that show untargeted metagenomic sequencing captures expected biological signals in wastewater samples.

**Figure 6.**
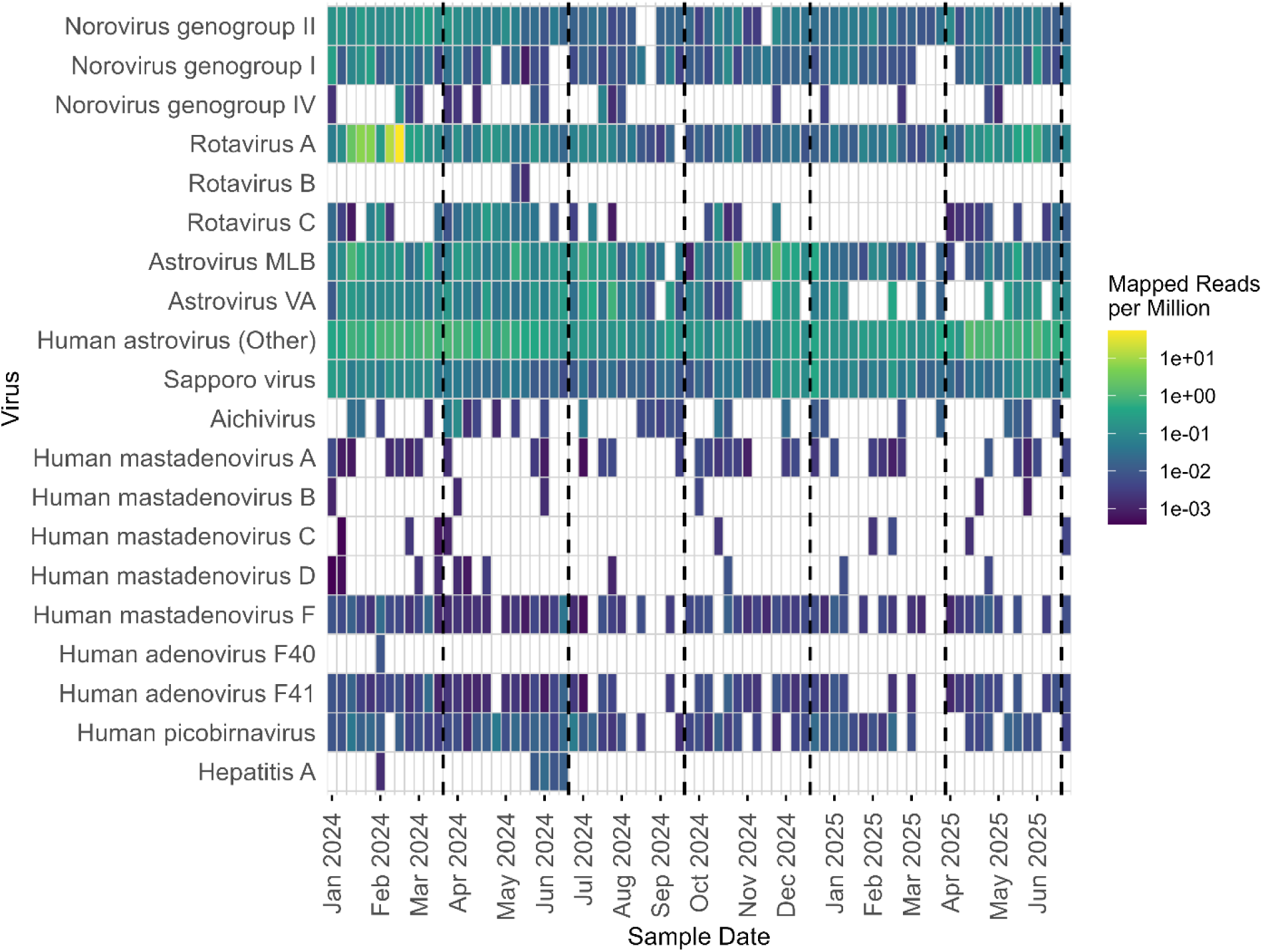
Normalized abundance (RPM) of *NVD* assigned virus reads associated with human gastrointestinal disease in Columbia MO wastewater from January 2024 to June 2025. Reads counts were derived by mapping raw sequences to NVD assembled contigs and counts were normalized to mapped reads per million prior to plotting as opposed to mapped reads per kilobase per million due to high level groupings of viruses with different genome lengths. A log_10_ color scale was used for visualization to enhance dynamic range and improve interpretability of variation across samples. Vertical dashed lines signify solstice and equinox dates to highlight season timeframes.

**Figure 7.**
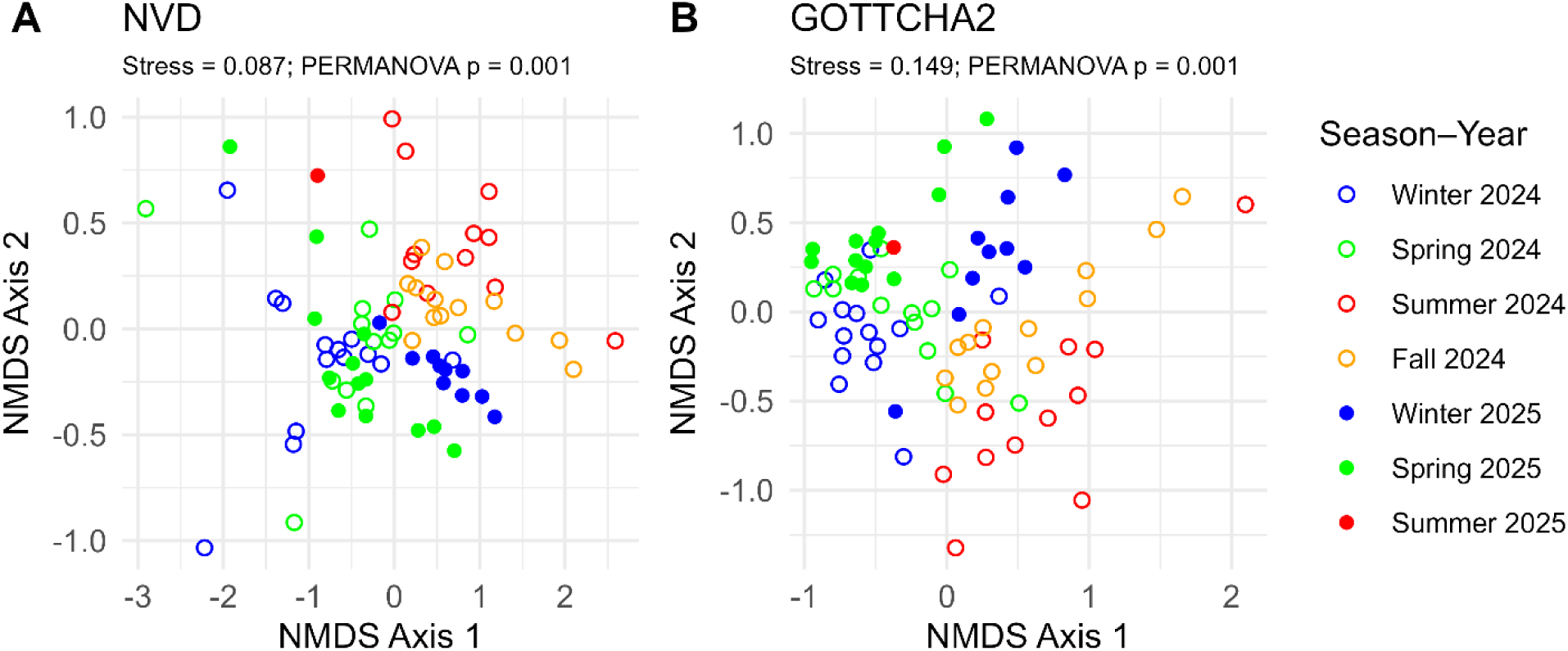
Sample clustering using non-metric multidimensional scaling of Bray-Curtis dissimilarity using human infecting virus counts derived from *NVD* and *GOTTCHA2*. Seasonal groupings were significant based on PERMANOVA results (p < 0.001 for both analysis).

## Discussion

This single-site longitudinal feasibility study demonstrates that an untargeted sequencing approach to detect human infecting viral sequences is possible from wastewater samples. By generating approximately one billion reads per week, we were able to detect a wide array of viruses, across a range of hosts, with sufficient sensitivity to track temporal trends and detect low-abundant or clinically overlooked pathogens. A major challenge in generating this type of data is the high proportion of rRNA sequences in the final output. While these ribosomal signals dilute target signals, they also provide important contextual information about the microbial communities, potential hosts, and other species signatures in wastewater. Additionally, removing rRNA during sample preparation introduces risks of removing target sequences. Given the low rate of classification of non-rRNA sequences using standard databases, there is a demand to generate a database of wastewater associated sequences for classifying future samples. While low classification rates in this study are consistent with previous work, Guajardo-Leiva, S. et al have recently shown that family level resolution is currently possible [24, 26, 38].

The detection of influenza C virus demonstrate that minimally biased sequencing approaches can capture clinically overlooked pathogens in a time-sensitive manner. While newly emerging HPAI strains such as H5N1 are available on commercially available panels, such as the Twist Comprehensive Viral Research, this minimally biased sequencing method recovered enough H5N1 sequences to facilitate partial assembly of five genomic segments with sufficient sequence information to resolve the virus to the B3.13 lineage. Additionally, the correlation between metagenomic read counts, and dPCR suggest that this method provides similar trends to current methods already being used for surveillance. The consistent detection of clinically relevant viruses such as SARS-CoV-2, parechovirus A, rhinovirus A, norovirus genogroup II, norovirus genogroup I, rotavirus A, astroviruses, Sapporo virus, human mastadenovirus F, and human picobirnavirus across all seasons strengthens the value of metagenomics for tracking endemic viral transmission in wastewater. Seasonal increases in virus abundance were observed for several of these pathogens, such as influenza virus, enterovirus A, and rotavirus A during colder months, which is consistent with established clinical trends. The similar human virus trends observed by multiple different taxonomic assignment methods between seasons reinforces the observations made in this study that are reflective of a wastewater dataset that follows traditionally expected temporal trends [2]

Beyond human health, the wide breadth of other detectable species emphasizes the expanded utility of metagenomic surveillance. We selected *GOTTCHA2* for definitive unambiguous characterization because this method is well suited for highly heterogeneous samples such as wastewater that contain a wide range of organisms such as viruses, bacteria, archaea, fungi, plants, and animals. The low classification rates using this method are a direct result of the fact that the reference database contains only unique kmers, and therefore the percentage classified is an underestimation of the total percentage of classifiable sequences. Other classifiers, such as *Kraken2* using the PlusPFP database, have potential to classify 93-99% of rRNA sequences or 14-55% of non-rRNA sequences across all taxonomy levels (data not shown). However, because that method contains non-unique kmers in the reference database, classification is less accurate. This ambiguity led to the decision to use *GOTTCHA2* for baseline characterization in this study due to its low false positive rate.

The detection of highly abundant plant viruses such as ToBRFV and TMV highlights this expanded utility in detecting unexpected sequences. While these viruses have geographic distributions that overlap with Missouri, it is likely that these viruses are introduced to wastewater through consumption of crops within *Solanaceae*, such as tomato and peppers, or suggest disease outbreaks in nearby plant communities, and further highlights the utility of an expanded breadth of detectable outbreaks when using metagenomic approaches to detect and identify viruses. While this study provides evidence that ToBRFV an optimal virus for use as an endogenous control in some locations, since ToBRFV detection is dependent on food sources, it may not be appropriate in all parts of the world with different diets and supply chains [26]. In addition to ToBRFV, the detection of plant viruses in this study is consistent with previous work showing high abundance of plant viruses in wastewater [23]. These results, in combination with human viral detection, demonstrate that deep metagenomic sequencing can support timely, minimally biased, and ecologically broad surveillance that supports the initiatives of CDC’s One Health. Heatmaps, similar to **Figures 4-6**, are publicly available so that trends can be followed in real time during the continuation of this study (https://lungfish-science.github.io/wastewater-dashboard/). The dataset produced here provides a starting point for other research efforts to begin development of new bioinformatic workflows to further improve the detection of impactful nucleic acid signatures in deep data.

As sequencing costs continue to decline and analytical workflows improve, these findings could support broader implementation of this approach as a scalable solution for public health preparedness and environmental monitoring. However, at current prices this sequencing approach may not be cost effective for all surveillance programs. This study is also limited due to single-site geographic representation. Additional sampling at new locations is required to determine if these findings can be generalized, and to fully evaluate scalability of this ultra deep sequencing method. Another limitation of this study is the evaluation of only non-solid fractions of wastewater. While it is known that some viruses are enriched in the solid fraction, we have not found an efficient method of separating viral material from solids that is sufficiently pure for untargeted metagenomics. Evaluation of the supernatant allowed us to utilize a PEG concentration method that adequately concentrated viruses without the use of rRNA depletion from samples. While these limitations restrict generalization of our findings, this study is a first look at how ultra deep sequencing at this depth can be applied to wastewater surveillance programs. Lastly, the data generated in this study can be analyzed many different ways, and analyses presented here should be viewed as one of several possible perspectives rather than a complete characterization.

## Acknowledgments

Authors would like to thank the University of Missouri Genomics Core for technical support in weekly sequencing efforts and optimization of library preparation protocols. Additionally, authors would like to thank the Research Support Solutions in the Division of IT at the University of Missouri and the Center for High Throughput Computing at the University of Wisconsin-Madison for access to high performance computing infrastructure (Columbia MO DOI: https://doi.org/10.32469/10355/97710). We would also like to thank Tami Hansen and the wastewater treatment plant in Columbia, MO for weekly sample collection. This research was funded by Inkfish, and by Coefficient Giving through a grant to SecureBio.

## Author Contributions

Clayton Rushford, Data curation, Formal analysis, Investigation, Methodology, Project Administration, Software, Supervision, Validation, Visualization, Writing – original draft, Writing – review and editing |Devon Gregory, Data curation, Formal analysis, Investigation, Methodology, Software, Validation, Visualization, Writing – review and editing | Emma Copen, Data Curation, Investigation, Methodology, Resources, Writing – review and editing | Jonathan Naydenov, Methodology | Alejandro Tovar-Mendez, Methodology, Writing – review and editing | Po-E Li, Data curation, Formal analysis, Software | Rose S. Kantor, Data curation, Methodology, Validation, Writing – review and editing | Migun Shakya, Data curation, Methodology, Validation, Writing – review and editing | Nelson Ruth, Software | Patrick S. G. Chain, Software | Hsinyeh Hsieh, Investigation, Methodology, Resources, Writing – Review & Editing | Torin Hunter, Investigation, Methodology, Writing – Review & Editing | Josh Kome, Software | Matt Frank, Investigation | Terri Lyddon, Resources | Jeff Kaufman, Conceptualization, Funding Acquisition, Methodology, Project administration, Writing – Review & Editing | David H. O’Connor, Conceptualization, Data Curation, Formal Analysis, Funding Acquisition, Methodology, Resources, Software, Validation, Visualization, Writing – Original Draft, Writing – Review & Editing | Marc C. Johnson, Conceptualization, Funding Acquisition, Methodology, Project Administration, Resources, Supervision, Validation, Writing – Original Draft, Writing – Review & Editing.

## Data Availability

Sequence data has been uploaded to the NCBI Sequence Read Archive under accession number PRJNA1247874. The workflow for identifying human-infecting virus sequences with high confidence is available at https://github.com/dhoconno/nvd/. The workflow for *GOTTCHA2* analysis, and subsequent visualization of *NVD* and *GOTTCHA2* results are available at https://github.com/ClaytonRushford/Columbia_MO_viral_surveillance_2024-2025/.

## Supplemental material

Table S1: Breakdown of treatments in each sample and sequencing platform used.

Table S2: Host annotation for *GOTTCHA2* detected phyla.

## Funding

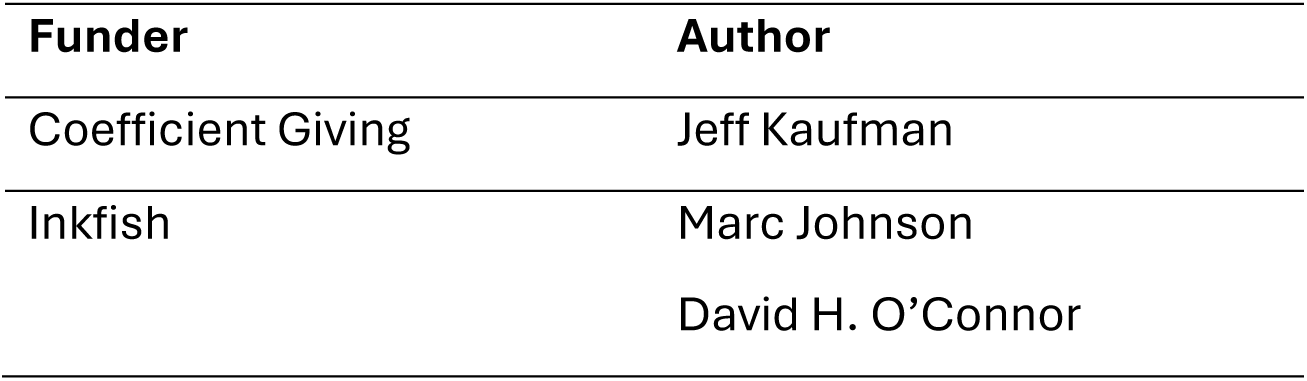

---

**Supplemental Figure 1.**
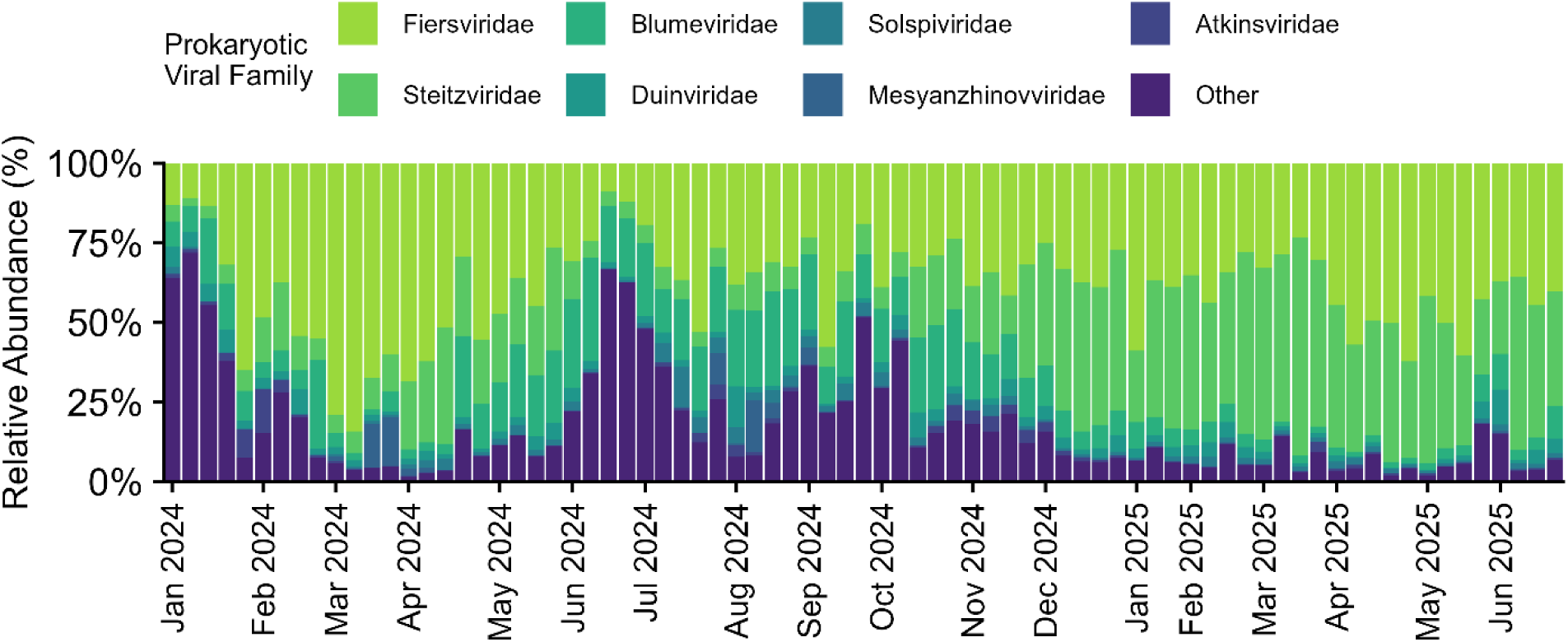
Normalized abundance (RPM) of *GOTTCHA2* classified viral families with prokaryotic hosts.

**Supplemental Figure 2.**
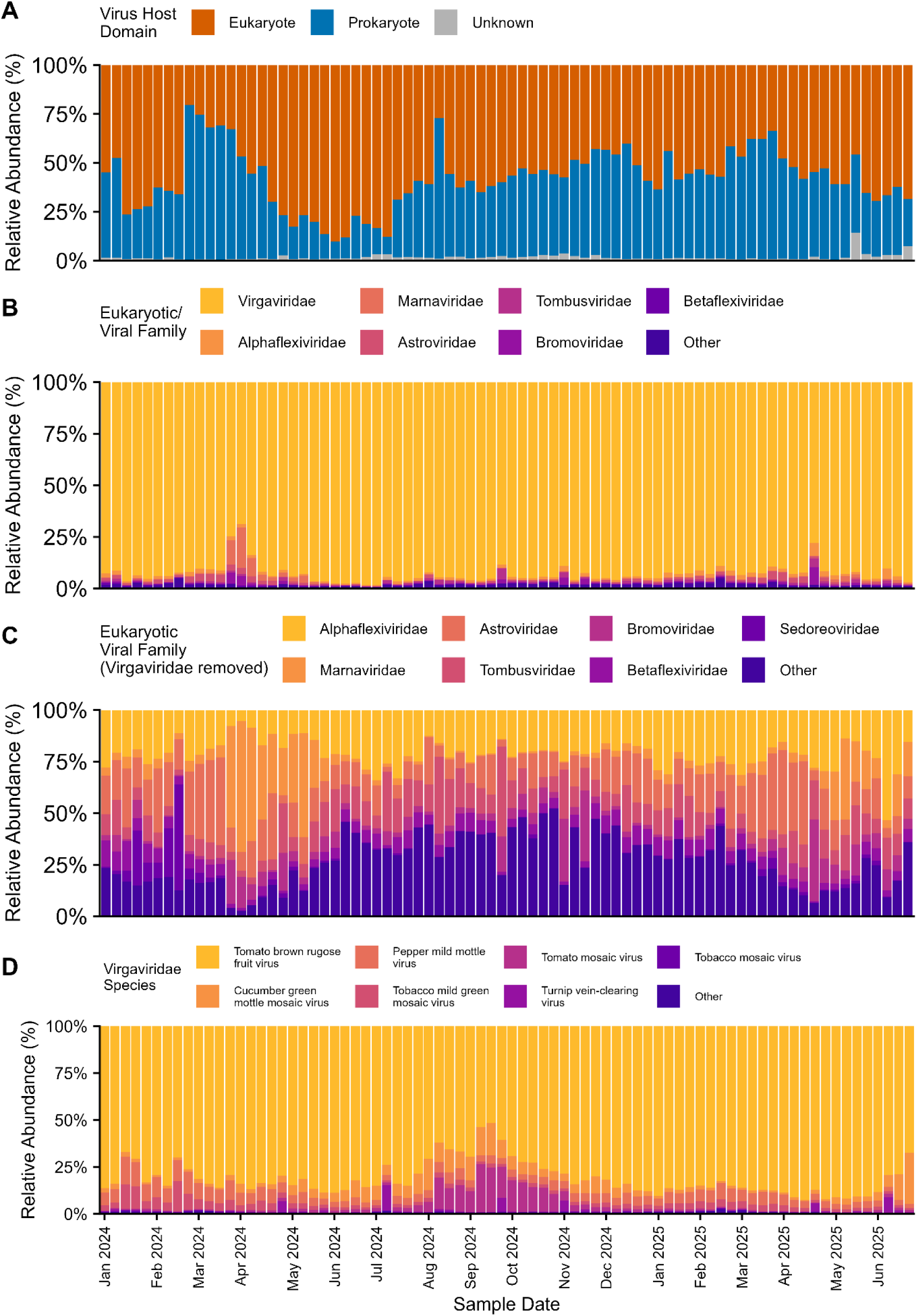
Normalized abundance (RPM) of *Kraken2* classified viral reads out of total classified viral reads, colored by host domain based on manual annotation (A). Normalized abundance (RPM) of the seven most abundant eukaryotic infecting virus families (B). Normalized abundance (RPM) of the seven most abundant families within eukaryotic viruses after removal of the family *Virgaviridae* (C). Breakdown of the most abundant species within the family *Virgaviridae* (D).

**Supplemental Figure 3.**
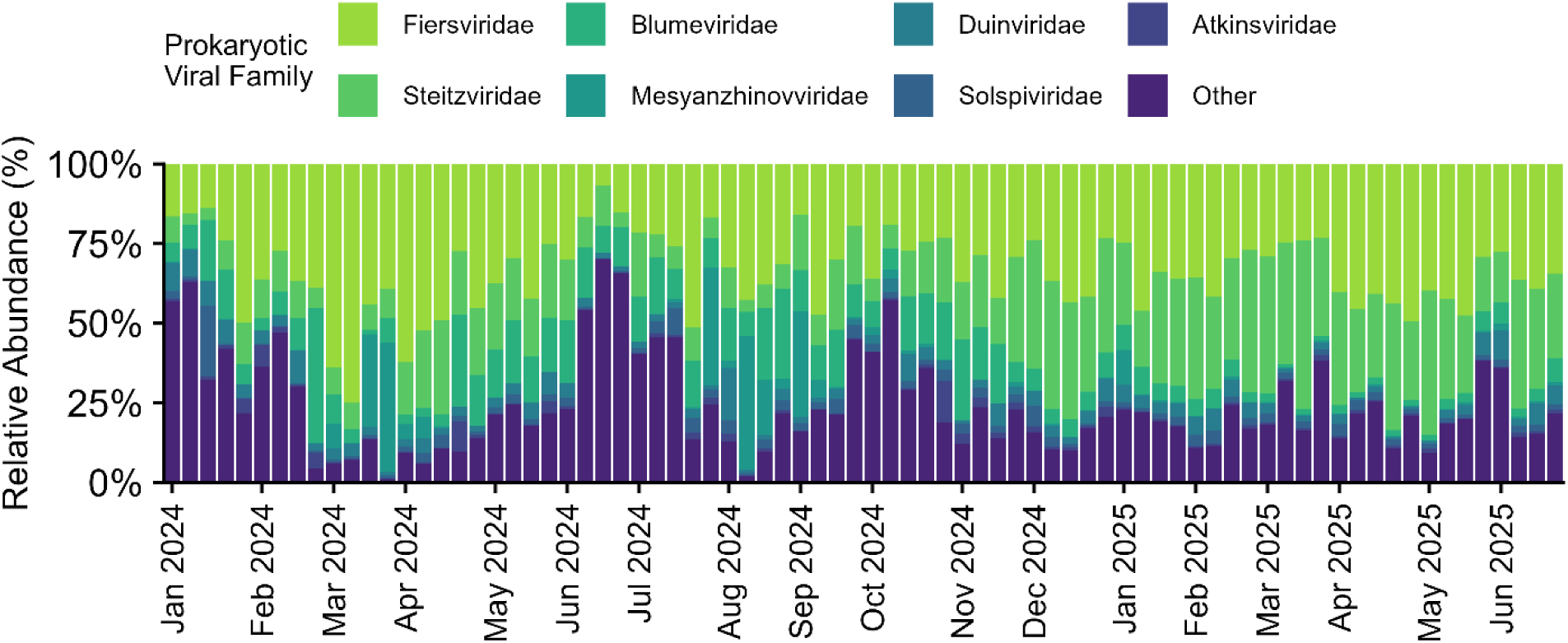
Normalized abundance (RPM) of *Kraken2* classified viral families with prokaryotic hosts.

**Supplemental Figure 4.**
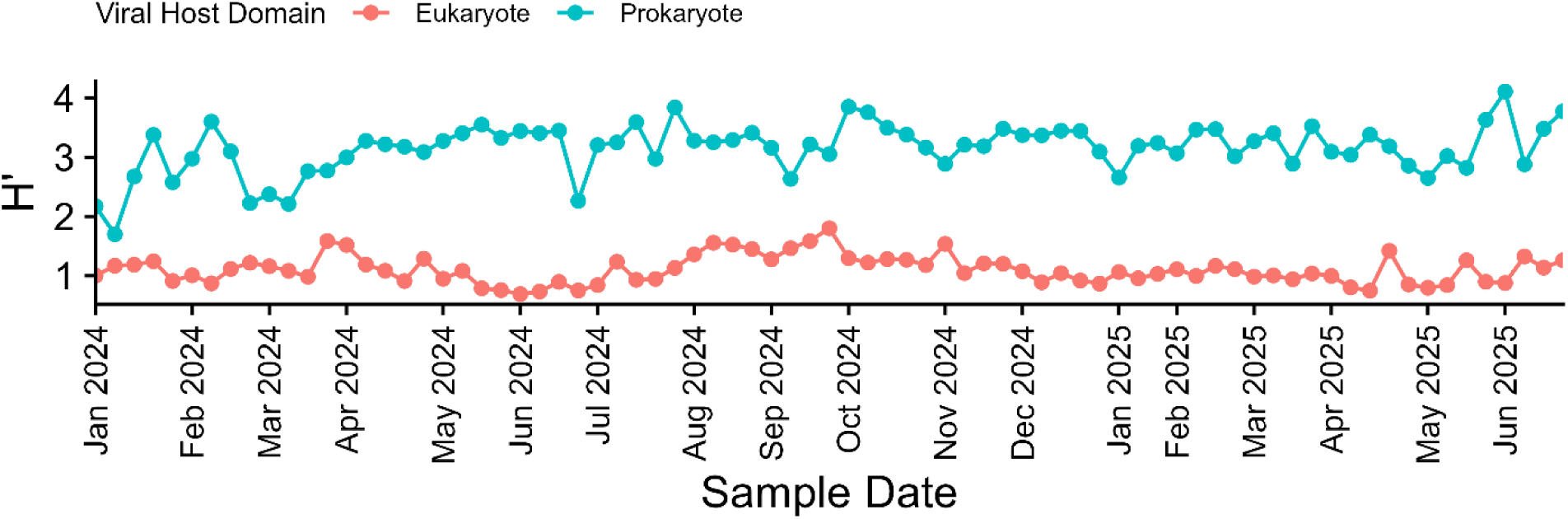
Shannon-Wiener diversity index values for *GOTTCHA2* assigned prokaryotic infecting (Blue) and eukaryotic infecting (Orange) virus groups. H′ was analyzed on the species level.

**Supplemental Figure 5.**
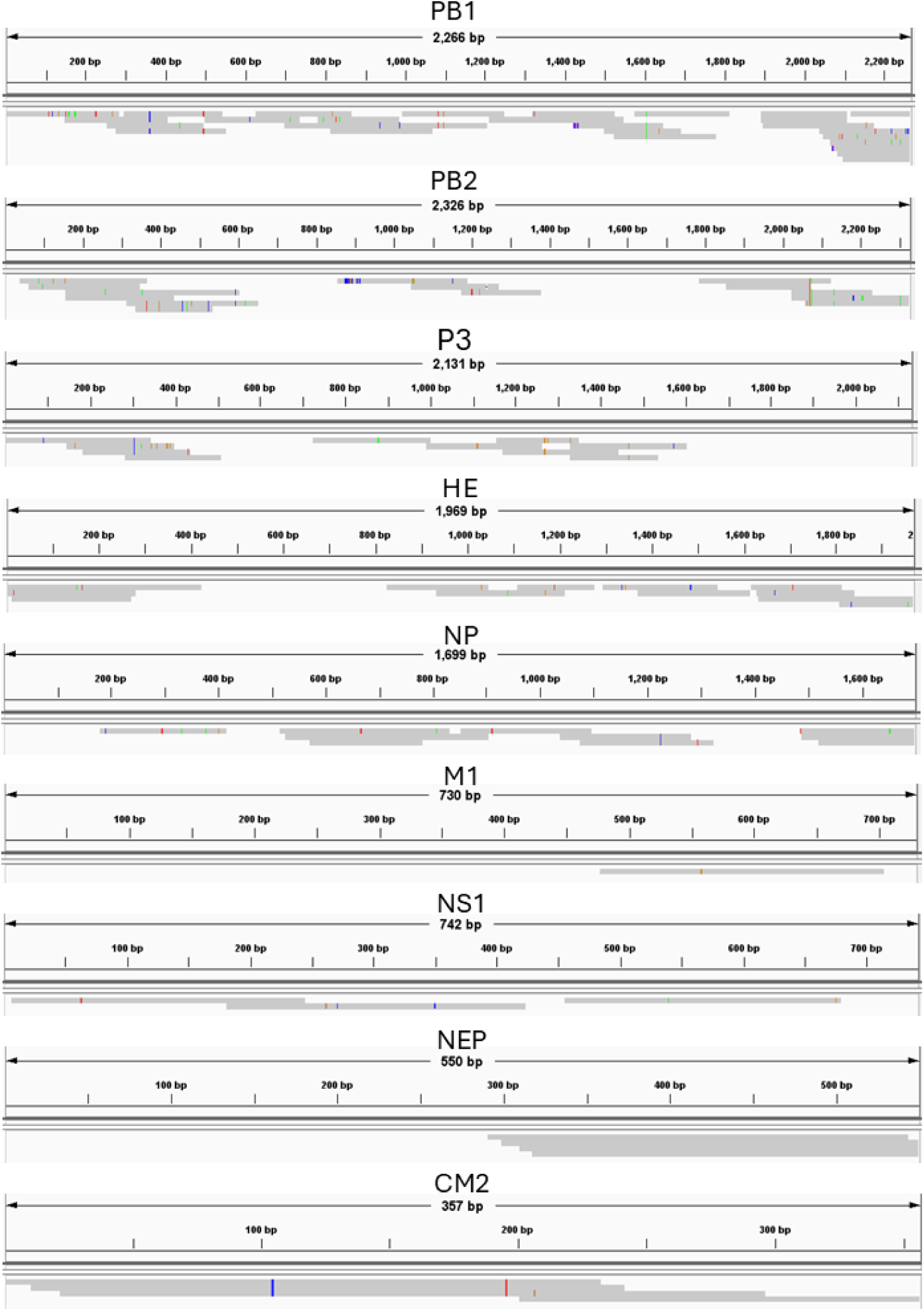
Alignment of NVD derived Influenza C assemblies mapped to coding sequences of Influenza C virus (C/Sri Lanka/68/2023) throughout the study period. Alignments were visualized on IGV, and images were cropped to only display the reference and coverage of the assembled contigs.

**Supplemental Figure 6.**
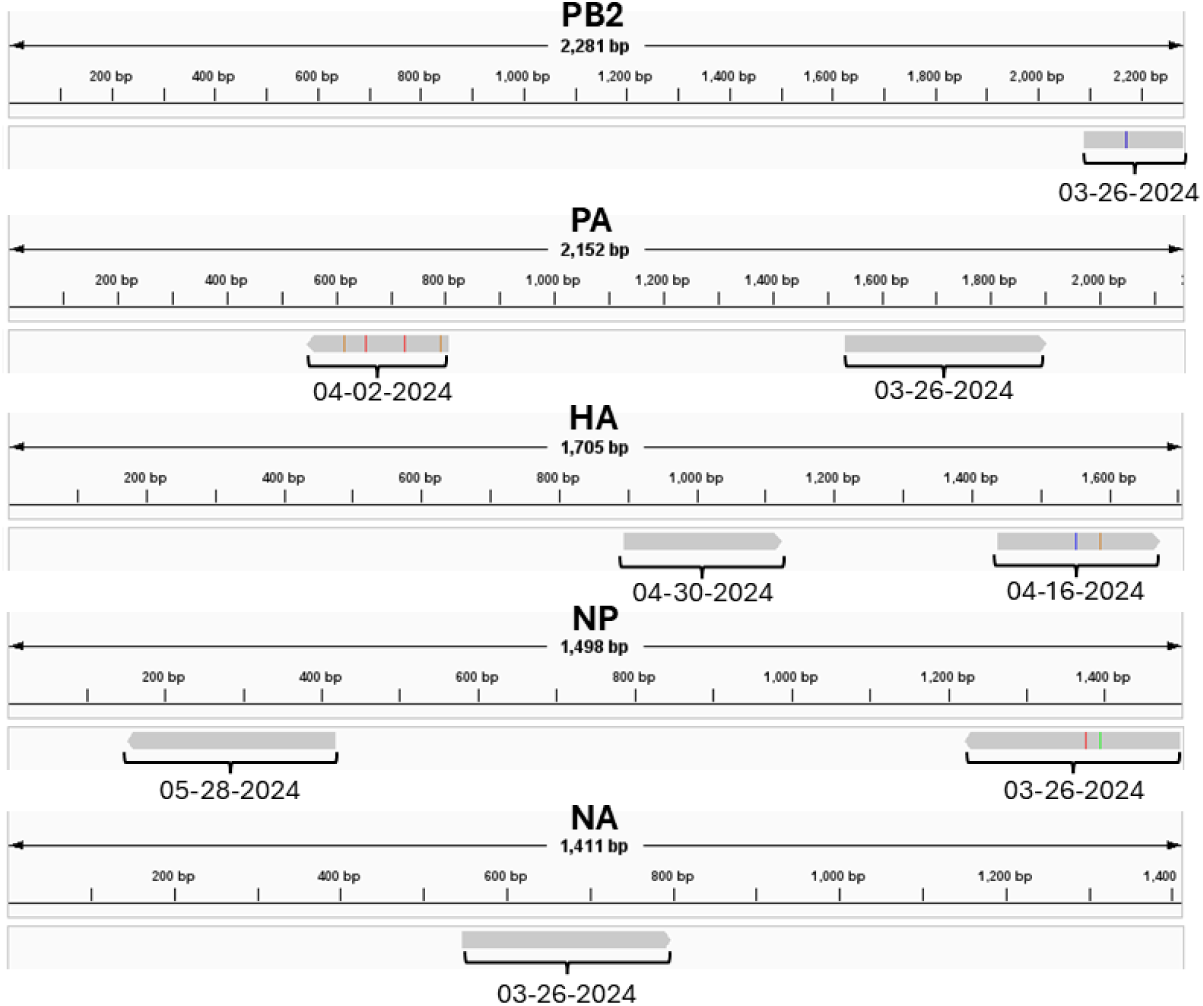
Alignment of NVD derived H5N1 assemblies mapped to Influenza A virus (A/Grackle/Texas/USL_047/2024(H5N1)). Alignments were visualized on IGV, and images were cropped to only display the reference and coverage of the assembled contigs. Sample dates associated with each contig are annotated in mm-dd-yyyy format. One contig H5N1 contig (218bp, BLAST validated as Influenza A virus (A/cat/ID/24-015315-001-original/2024(H5N1)) segment 3 polymerase) was filtered out from this alignment.

## References

1. Grassly, N.C., A.G. Shaw, and M. Owusu, Global wastewater surveillance for pathogens with pandemic potential: opportunities and challenges. The Lancet Microbe, 2024.

2. Smith, M.F., et al., Seasonality of respiratory, enteric, and urinary viruses revealed by wastewater genomic surveillance. Msphere, 2024. 9(5): p. e00105–24.

3. Kilaru, P., et al., Wastewater surveillance for infectious disease: a systematic review. American journal of epidemiology, 2023. 192(2): p. 305–322.

4. Majumdar, M., et al., Detection of Enterovirus D68 in wastewater samples from the United Kingdom during outbreaks reported globally between 2015 and 2018. BioRxiv, 2019: p. 738948.

5. Jahn, K., et al., Early detection and surveillance of SARS-CoV-2 genomic variants in wastewater using COJAC. Nature Microbiology, 2022. 7(8): p. 1151–1160.

6. Kim, N.-Y., et al., Wastewater Knows Pathogen Spread: Analysis of Residential Wastewater for Infectious Microorganisms including SARS-CoV-2. Infection & Chemotherapy, 2023. 55(2): p. 214.

7. Kantor, R.S. and M. Jiang, Considerations and Opportunities for Probe Capture Enrichment Sequencing of Emerging Viruses from Wastewater. Environmental Science & Technology, 2024. 58(19): p. 8161–8168.

8. Santos, A.F., et al., Wastewater Metavirome Diversity: Exploring Replicate Inconsistencies and Bioinformatic Tool Disparities. International Journal of Environmental Research and Public Health, 2025. 22(5): p. 707.

9. Pogka, V., et al., Targeted virome sequencing enhances unbiased detection and genome assembly of known and emerging viruses—the example of SARS-CoV-2. Viruses, 2022. 14(6): p. 1272.

10. McCall, C., et al., Targeted metagenomic sequencing for detection of vertebrate viruses in wastewater for public health surveillance. ACS ES&T Water, 2023. 3(9): p. 2955–2965.

11. Tisza, M., et al., Wastewater sequencing reveals community and variant dynamics of the collective human virome. Nature Communications, 2023. 14(1): p. 6878.

12. Zulli, A., et al., Probe-Based Enrichment Sequencing Applied to Wastewater Surveillance Accurately Tracks Multiple Viral Respiratory Outbreaks. ACS ES&T Water, 2025.

13. Javornik Cregeen, S., et al., Sequencing-Based Detection of Measles in Wastewater: Texas, January 2025. American Journal of Public Health, 2025(0): p. e1–e5.

14. Clark, J.R., et al., Wastewater pandemic preparedness: Toward an end-to-end pathogen monitoring program. Frontiers in Public Health, 2023. 11: p. 1137881.

15. Thornton, M., et al., Comparative wastewater virome analysis with different enrichment methods. Water Research, 2025: p. 123985.

16. Liu, S., et al., Analysis of metagenomic data. Nature Reviews Methods Primers, 2025. 5(1): p. 5.

17. Wetterstrand, K. DNA Sequencing Costs: Data from the NHGRI Genome Sequencing Program.

18. Rajput, V., et al., Metagenomics based longitudinal monitoring of antibiotic resistome and microbiome in the inlets of wastewater treatment plants in an Indian megacity. Science of The Total Environment, 2025. 986: p. 179691.

19. Becsei, Á., et al., Time-series sewage metagenomics distinguishes seasonal, human-derived and environmental microbial communities potentially allowing source-attributed surveillance. Nature communications, 2024. 15(1): p. 7551.

20. Child, H.T., et al., Comparison of metagenomic and targeted methods for sequencing human pathogenic viruses from wastewater. MBio, 2023. 14(6): p. e01468–23.

21. Li, Y., et al., A broad wastewater screening and clinical data surveillance for virus-related diseases in the metropolitan Detroit area in Michigan. Human Genomics, 2024. 18(1): p. 14.

22. Cantalupo, P.G., et al., Raw sewage harbors diverse viral populations. MBio, 2011. 2(5): p. 10.1128/mbio.00180-11.

23. Nieuwenhuijse, D.F., et al., Setting a baseline for global urban virome surveillance in sewage. Scientific reports, 2020. 10(1): p. 13748.

24. Guajardo-Leiva, S., J. Chnaiderman, A. Gaggero, and B. Díez, Metagenomic insights into the sewage RNA virosphere of a large city. Viruses, 2020. 12(9): p. 1050.

25. Gulino, K., et al., Initial mapping of the New York City wastewater virome. Msystems, 2020. 5(3): p. 10.1128/msystems.00876-19.

26. Guajardo-Leiva, S., et al., From sewage to genomes: Expanding our understanding of the urban and semi-urban wastewater RNA virome. Environmental Research, 2025. 276: p. 121509.

27. Wyler, E., et al., Pathogen dynamics and discovery of novel viruses and enzymes by deep nucleic acid sequencing of wastewater. Environment International, 2024. 190: p. 108875.

28. Kitajima, M., H.P. Sassi, and J.R. Torrey, Pepper mild mottle virus as a water quality indicator. NPJ Clean Water, 2018. 1(1): p. 19.

29. Bushnell, B. BBmap. 2025 2025-06-05 [cited 2025 6-20]; Available from: https://sourceforge.net/projects/bbmap/.

30. Freitas, T.A.K., P.-E. Li, M.B. Scholz, and P.S. Chain, Accurate read-based metagenome characterization using a hierarchical suite of unique signatures. Nucleic acids research, 2015. 43(10): p. e69–e69.

31. Robinson, C.A., et al., Defining biological and biophysical properties of SARS-CoV-2 genetic material in wastewater. Science of The Total Environment, 2022. 807: p. 150786.

32. Russell, E., A. Yang, S. Tardrew, and M.G. Ison, Parainfluenza virus in hospitalized adults: a 7-year retrospective study. Clinical Infectious Diseases, 2019. 68(2): p. 298–305.

33. Control, C.f.D. and Prevention, Influenza activity in the United States during the 2023–2024 season and composition of the 2024–2025 influenza vaccine. 2024.

34. Grizer, C.S., K. Messacar, and J.J. Mattapallil, Enterovirus-D68–a reemerging non-polio enterovirus that causes severe respiratory and neurological disease in children. Frontiers in Virology, 2024. 4: p. 1328457.

35. Dhar, D., et al., Emergence of Parechovirus-A5 central nervous system infections in children from Kansas City, Missouri, USA. Journal of Clinical Virology, 2025: p. 105835.

36. Campbell, A., K. Brizuela, and S.S. Lakdawala, mGem: Transmission and exposure risks of dairy cow H5N1 influenza virus. MBio, 2025. 16(3): p. e02944–24.

37. Tisza, M.J., et al., Virome Sequencing Detects H5N1 Avian Influenza in Wastewater in Ten Cities. The New England journal of medicine, 2024. 391(12): p. 1157.

38. Tamaki, H., et al., Metagenomic analysis of DNA viruses in a wastewater treatment plant in tropical climate. Environmental microbiology, 2012. 14(2): p. 441–452.

